# Microbiologic Spectrum and Antimicrobial Resistance in Febrile Neutropenia: Experience from an Eastern India Hematology Centre

**DOI:** 10.1101/2025.03.22.25324438

**Authors:** Prakas Kumar Mondal, Christy Varghese John, Rayhan Sekh, Praveen Kumar, Shuvra Neel Baul, Tuphan Kanti Dolai

## Abstract

**Background:** Febrile neutropenia (FN), a life-threatening complication of chemotherapy, is a medical emergency requiring prompt antibiotic therapy. The epidemiology of causative pathogens is evolving with a shift towards multidrug-resistant (MDR) strains. This study aimed to evaluate the microbiologic spectrum and antimicrobial resistance patterns in FN patients at a hematology center in Eastern India.

**Methods:** A retrospective study was conducted on FN patients admitted between July 2022 and July 2024. Clinical and demographic data, along with microbiological isolates and their antimicrobial susceptibility, were analyzed. Samples were evaluated from blood, respiratory, urinary, skin and mucocutaneous sites. Antimicrobial susceptibility testing was performed using Vitek 2. Statistical analyses, including multinominal logistic regression, were performed to identify factors associated with MDR infections.

**Results:** Among the 1,604 FN episodes analyzed, the most common underlying hematologic malignancies were acute myeloid leukemia (46.2%) and acute lymphoblastic leukemia (34.3%). Blood culture positivity was 39%, with Gram-negative bacteria (GNB) (67.8%) predominating over Gram-positive bacteria (32.1%). The most frequently isolated GNB were *Klebsiella oxytoca* (12.4%), *Klebsiella pneumoniae* (8.5%), and *Escherichia coli* (7.3%). MDR isolates accounted for 30% of all pathogens, with significant resistance observed to fluoroquinolones (Ciprofloxacin: 57.8%) and carbapenems (Meropenem: 42.8%). MDR isolates of concern included carbapenem-resistant *Acinetobacter baumannii* (1.8%), carbapenem-resistant Enterobacterales (3.3%), and *Candida auris* (0.6%).

**Conclusion:** This study necessitates ongoing surveillance and antimicrobial stewardship among FN patients in view of emerging MDR strains. The current predominance of Enterobacterales suggests a need to re-evaluate empirical antibiotic choices.

Optimizing treatment strategies with newer β-lactam/β-lactamase inhibitors and targeted antimicrobial stewardship programs is essential in resource-limited settings.

## Introduction

Febrile neutropenia (FN) continues to be a major dose-limiting toxicity of anticancer treatment, especially in systemic chemotherapy for hematologic malignancies. FN often leads to dose reductions and suboptimal treatment regimens with the potential for compromised clinical outcomes in otherwise responsive malignancies(1). Its impact has been addressed with the help of myeloid growth factors, predictive risk-based scoring systems and prophylactic antibiotics. However, FN remains a medical emergency, prompting immediate evaluation and empirical treatment with broad-spectrum antimicrobials(2).

Neutropenia is defined by the sum of the circulating segmented and band neutrophils or the absolute neutrophil count (ANC). It is identified by an ANC below 1.5 × 10^9^/L. However, bacterial infection risk rises significantly with an ANC below 0.5 × 10^9^/L and is termed severe neutropenia(1).

Recent studies suggest that fever is a non-specific response to multiple insults, including infection. It may be the manifestation of a deranged host response to tissue damage at epithelial and mucosal junctions, which are inhabited by the body’s microbiome. This predisposes to opportunistic pathogens inhabiting the skin and alimentary tract(3).

Advances in FN management have been attributed to effective empiric antibiotic regimens. Frequent epidemiologic studies in various centres demonstrated the spectrum of bacterial pathogens isolated in FN patients(4). Studies from Western centres documented a shift from gram-negative species in the 1960s to more gram-positive species in the 1990s(1,5). Various studies demonstrated the mortality benefit of antipseudomonal coverage in patients with acute myeloid leukemia (AML)(1). At present, the most common species isolated from blood are coagulase-negative staphylococci(5). Fever remains unexplained in 30-50% of FN, and bloodstream infections are detected in 30%(1).

Emerging antimicrobial resistance mandates the judicious use of antimicrobial agents and surveillance to optimize existing empiric antibiotic regimens. However, microbiologically identified infections account for 20-25%. Recent surveys in the US performed in 2016-19 validate the adequacy of cefepime and piperacillin-tazobactam as empiric antibiotic therapy for high-risk hematologic malignancy patients experiencing their first episode of FN(6). The microbiological isolates were divided equally among aerobic Gram-negative and Gram-positive organisms. Enterobacteriaceae and *P. aeruginosa* made up 95% of Gram-negative BSI organisms. The Gram-positive organisms included viridans group streptococci (49%), followed by *Staphylococcus aureus* (17%) and coagulase-negative staphylococci (14%).(6)

## Materials and Methods

A retrospective study involving all patients of hematologic diseases with febrile neutropenia admitted to the Dept. of Hematology, Nil Ratan Sircar Hospital, between July 2022 and July 2024 was done.

The clinical and demographic data collected for each patient included sex, age, underlying hematologic disorder, and the absolute neutrophil count (ANC) at the onset of fever. The microbiological data gathered included the type of sample tested (bloodstream or otherwise), organisms isolated and their antimicrobial susceptibility patterns.

Two sets of blood samples were obtained from each patient. In patients with a central venous catheter (CVC), one set was obtained from a peripheral vein puncture and the other from a CVC. In patients without a CVC, two sets of blood samples were collected 30 minutes apart from two different peripheral venipuncture sites. Blood culture results were obtained using an automated blood culture system (BACTEC FX, Becton Dickinson, Sparks, MD, USA). The positive blood culture bottle was sent to Vitek 2 (bioMérieux, Hazelwood, MO, USA) for direct identification and antimicrobial susceptibility testing. The minimum inhibitory concentration values were classified according to the Clinical and Laboratory Standards (CLSI) guidelines, specifically the 34^th^ edition, published in 2024.

### Definitions

Neutropenia was defined as an ANC of <500 cells/mm^3^ or an ANC of <1000 cells/mm^3^ that was expected to fall to <500 cells/mm3. Different organisms isolated from a single patient ≥48 hours apart were considered separate FN episodes. Pathogens demonstrating an intermediate antimicrobial susceptibility were considered antimicrobial resistant.

Multi-drug resistant (MDR) bacteria were defined as isolates resistant to at least one agent in three or more different classes.

### Statistical analyses

The clinical and microbiological characteristics of the FN episodes are presented using counts and percentages for categorical variables and using medians and ranges for continuous variables. The number and nature of microbiological isolates are represented using counts and percentages. Multivariate logistic regression models were implemented to analyze risk factors for MDR isolates. A two-sided P-value of <0.05 was considered the threshold for statistical significance. All statistical analyses were performed using RStudio software (version 2024.12.0+467; Boston, MA, USA).

## Results

The clinical characteristics of the evaluated FN episodes are described in Table 1. A total of 1604 episodes were assessed, 64.7% of which occurred in men. The median age of presentation was 27 years. Patients with Acute Myeloid Leukemia (AML) accounted for 46.2%, followed by Acute Lymphoblastic Leukemia (ALL; 34.3%), Aplastic Anemia (AA; 7.2%) and Non-Hodgkin Lymphoma (NHL; 4.7%).

**Table 1:** Clinical characteristics of febrile neutropenia episodes.

| Characteristic | Total |
| --- | --- |
| **Demographics** |  |
| Male sex (M) | 1,038 (64.7%) |
| Age (years), median (range) | 27 (1-87) |
| **Hematologic Parameters** |  |
| Median ANC (IQR) | 500 (800) |
| **Hematologic Diseases** |  |
| Aplastic Anemia (AA) | 117 (7.2%) |
| Acute Lymphoblastic Leukemia (ALL) | 552 (34.3%) |
| Acute Myeloid Leukemia (AML) | 742 (46.2%) |
| Chronic Lymphocytic Leukemia (CLL) | 4 (0.2%) |
| Chronic Myeloid Leukemia (CML) | 25 (1.5%) |
| Hodgkin Lymphoma (HL) | 11 (0.7%) |
| Myelodysplastic Syndrome (MDS) | 20 (1.3%) |
| Multiple Myeloma (MM) | 58 (3.6%) |
| Non-Hodgkin Lymphoma (NHL) | 75 (4.7%) |

**Table 2:**
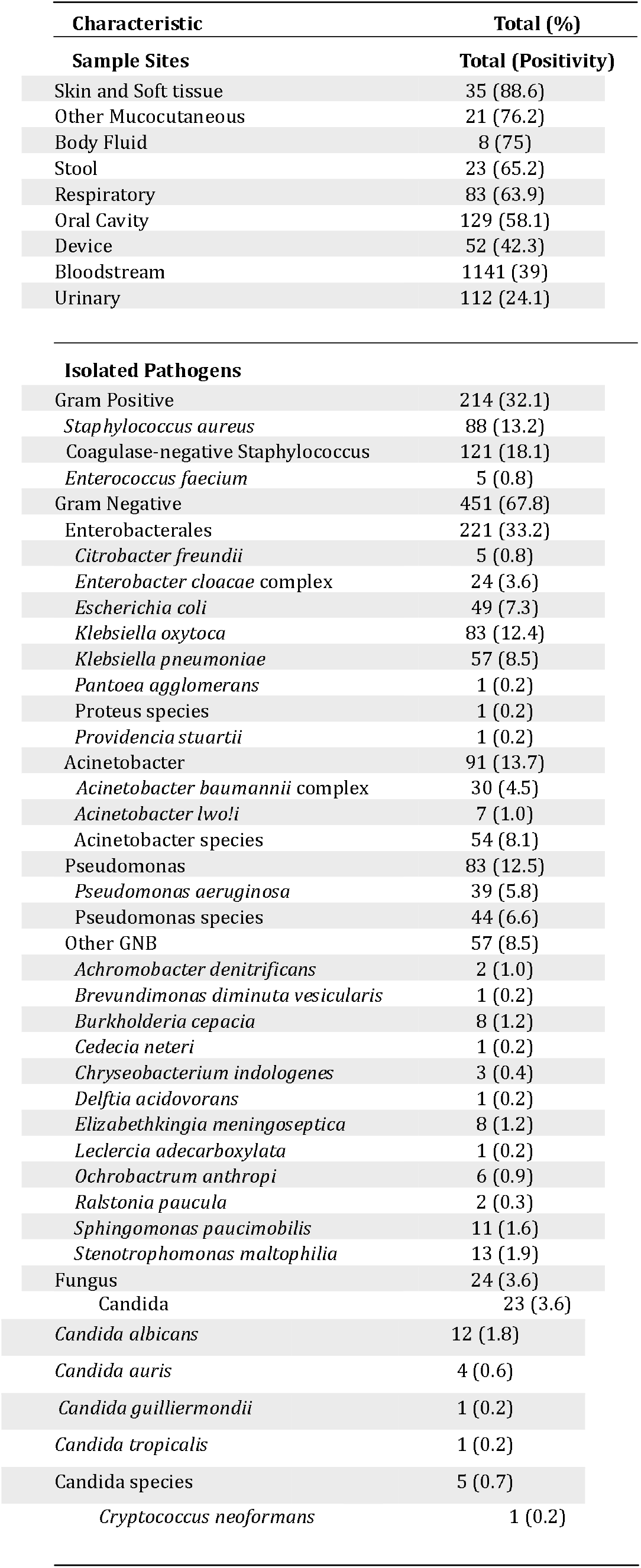
Microbiological characteristics of febrile neutropenia episodes.

Among the microbiological samples sent for evaluation, 71.1% were bloodstream samples, followed by oral cavity samples (8%) and urinary samples (6.9%).

Microorganisms were isolated in 41.4% (665) of all samples, with the highest among skin and soft tissue samples (88.6%), followed by samples from mucocutaneous sites other than the oral cavity (76.2%). The blood culture positivity rate was 39%.

Among the isolated microorganisms, Gram-negative bacteria accounted for 67.8% (451). Gram-positive bacteria accounted for 32.1% (214), while fungal species accounted for 3.6% (24) of all isolates.

Among the Gram-negative bacteria (GNB), species belonging to the order Enterobacterales (221, 33.2%), such as *Klebsiella oxytoca* (83, 12.4%), *Klebsiella pneumoniae* (57, 8.5%) and *Escherichia coli* (49, 7.3%) were predominant. It also included Acinetobacter species (91, 13.7%), Pseudomonas species (83, 12.5%) and other notable non-lactose fermenter species such as *Stenotrophomonas maltophilia* (13, 1.9%), *Burkholderia cepacia* (8, 1.2%) and *Elizabethkingia meningoseptica* (8, 1.2%).

The isolated fungal species were predominantly Candida species (23, 3.6%), notably *Candida albicans* (12, 1.8%) and *Candida auris* (4, 0.6%), and a single isolate of *Cryptococcus neoformans*.

The rates of antimicrobial resistance were high among all classes tested, especially Oxacillin (91.9%), Ceftazidime (71.9%) and Piperacillin/Tazobactam (53.2%) among Beta lactams. Among Carbapenems, the resistance rates were highest for Doripenem (54.2%), followed by Meropenem (42.8%), Imipenem (39.3%) and Ertapenem (36.4%).

Notably, resistance rates were high for Fluoroquinolones, especially Ciprofloxacin (57.8%) and Levofloxacin (47.2%), and Aminoglycosides such as Gentamicin (44.1%) and Tobramycin (36.8%). However, resistance rates were more favourable for Linezolid (10.6%), Tigecycline (8.6%) and Glycopeptides such as Vancomycin (18.5%) and Daptomycin (undetected).

The proportion of MDR isolates among all samples was 30%. The proportion of MDR isolates was maximal in the bloodstream (31%), followed by urinary (25.9%), mucocutaneous sites other than the oral cavity (25%) and respiratory samples (22.6%). 13.6% of the isolates sampled from intravascular devices were MDR.

Among the bacterial isolates, the proportion of MDR isolates was highest in Coagulase-negative staphylococci (CoNS) (45; 37.2%), followed by Pseudomonas species (29; 34.9%), Acinetobacter species (22; 24%), other non-lactose fermenting Gram-negative bacteria (21; 36.8%) and Enterobacterales (51; 23%). None of the fungal isolates demonstrated antifungal resistance.

Among antimicrobial-resistant pathogens of concern, the isolates identified included Carbapenem-resistant Acinetobacter baumanii (CRAb) (13; 1.8%), Carbapenem-resistant Enterobacterales (CRE) (23; 3.3%), *Stenotrophomonas maltophilia* (13; 1.9%), Methicillin-resistant *Staphylococcus aureus* (MRSA) (9; 1.3%), Difficult-to-treat Pseudomonas (5; 0.7%) and *Candida auris* (4; 0.6%).

### Risk factors of MDR Bloodstream Infection

The predictive factors associated with MDR infections, as evidenced by MDR isolates, are provided in Table 4. Multivariate analysis was done to assess the association of underlying hematologic disease, demographic factors and sampling site with the isolation of an MDR pathogen.

**Table 3:**
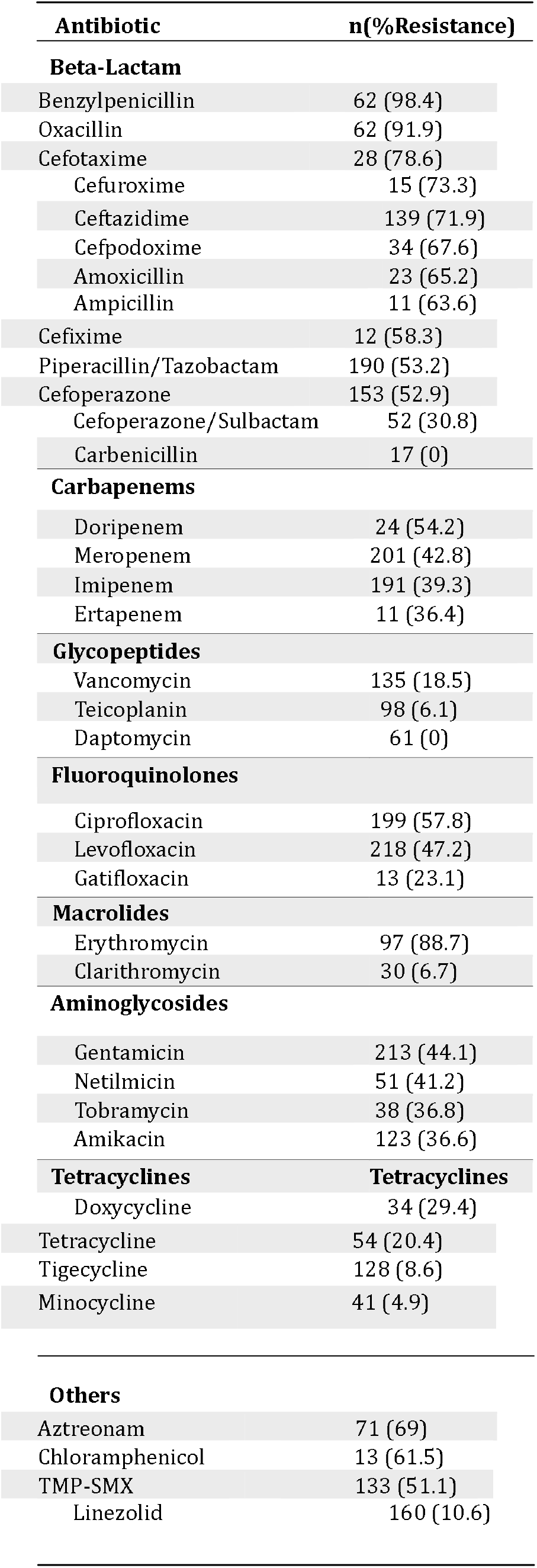
Antimicrobial resistance of isolates.

**Table 4:**
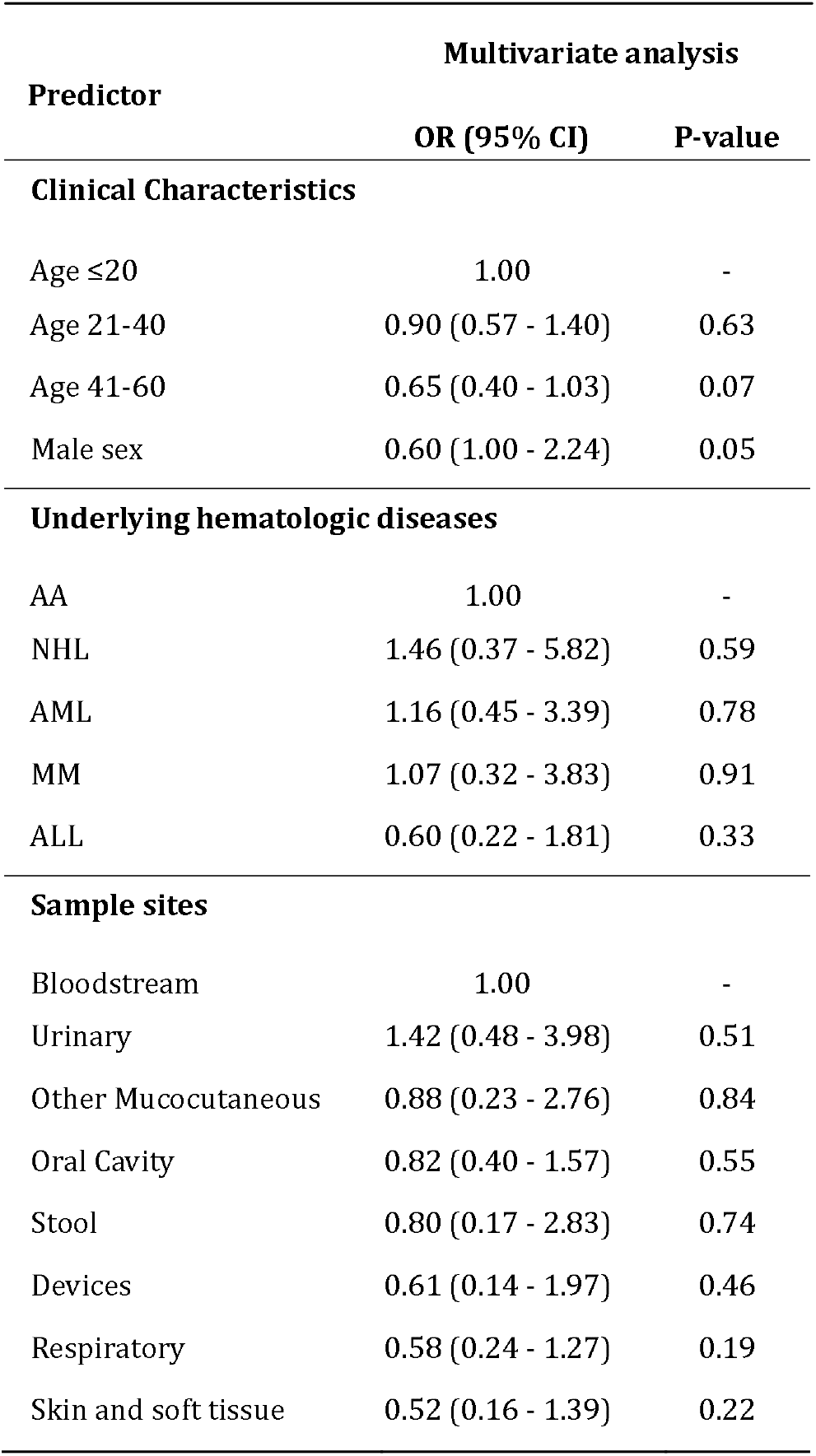
Factors associated with multidrug-resistant infections in FN patients.

**Figure 1:**
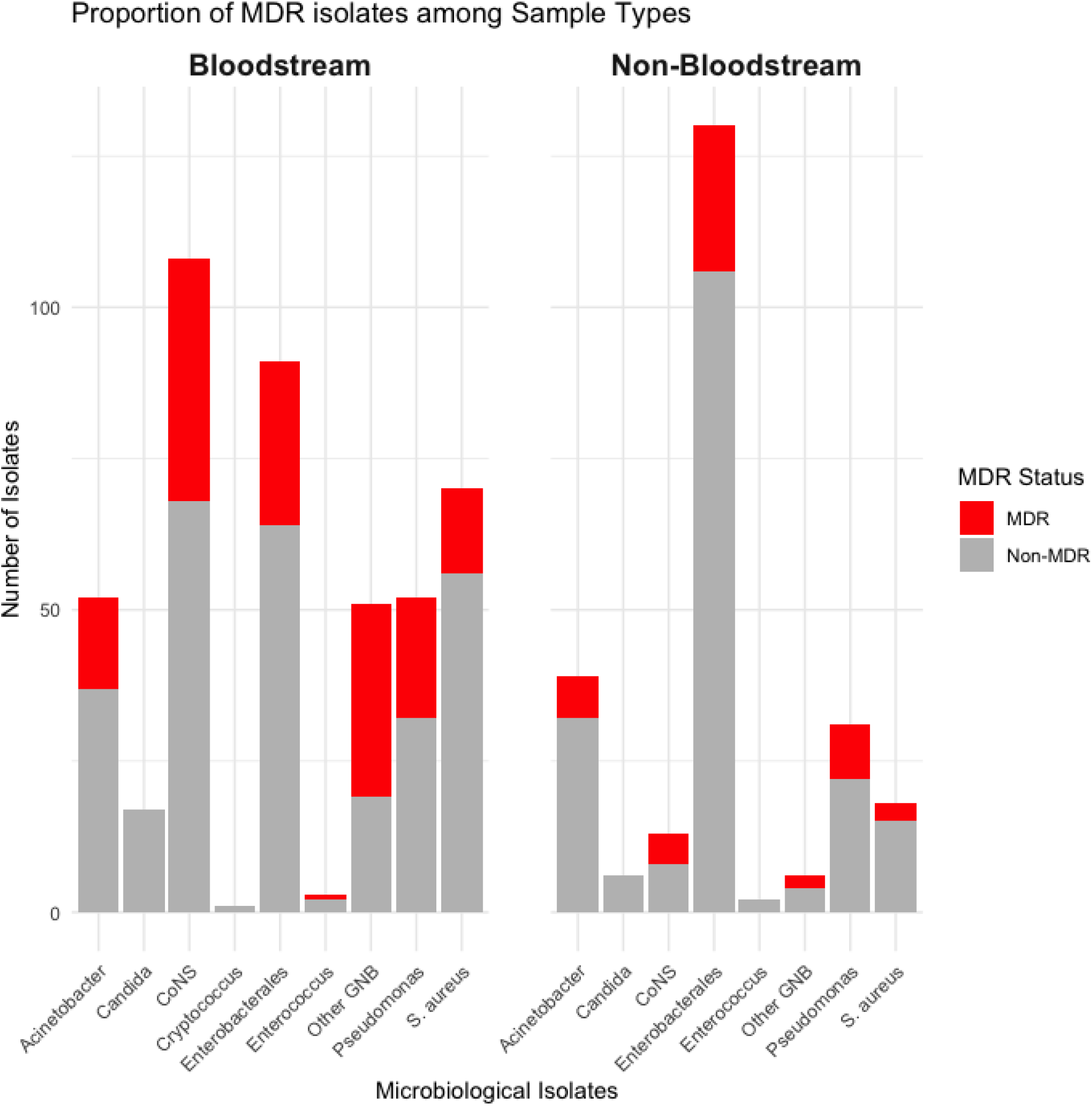
Proportion of MDR isolates among sample types.

**Figure 2:**
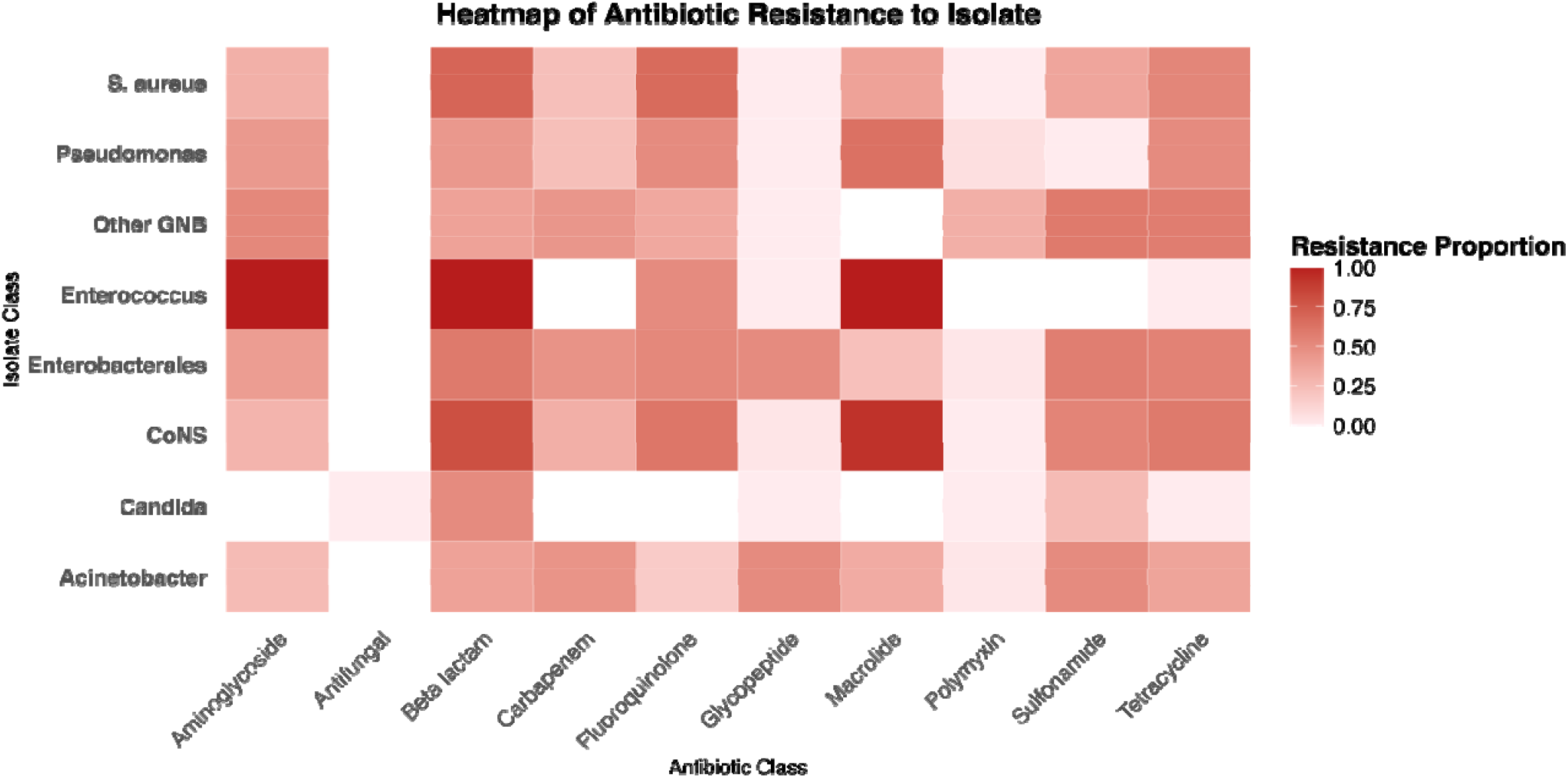
Antimicrobial resistance pattern across isolated pathogens.

**Figure 3:**
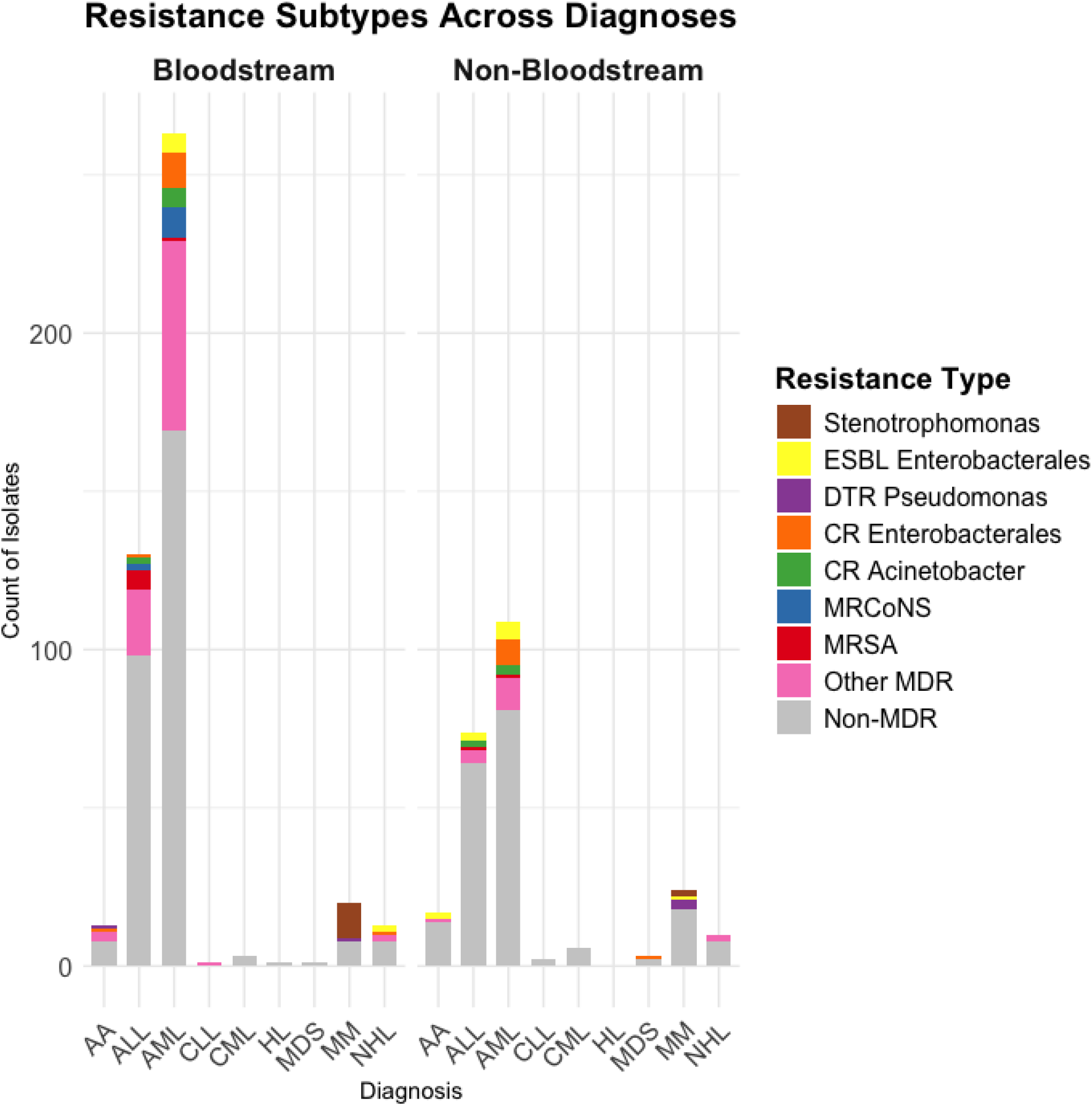
MDR isolates of concern among hematologic diseases.

## Discussion

Our single-centre microbiological surveillance study incorporates bloodstream infections with samples procured from other sites. The most isolated pathogens were Gram-negative bacilli, belonging to the order Enterobacterales namely *Klebsiella oxytoca* (12.4%), *Klebsiella pnuemoniae* (8.5%), and *Escherichia coli* (7.3%). However, among bloodstream isolates, the most isolated were Coagulase-negative staphylococci (24.3%), Enterobacterales (20.5%), namely and *Staphylococcus aureus* (15.7%). This is representative of the evolving nature of the commonly isolated pathogens and their resistance patterns in our institute over the past decade. Our current study highlights the prominence of Enterobacterales, namely Klebsiella species, over Pseudomonas as the predominant pathogenic isolate(7). This shift may be attributed to the widespread use of empirical antipseudomonal therapy and is witnessed in similar resource-limited settings(4).

The isolates from intravascular devices included *Staphylococcus aureus* (6, 27%), Acinetobacter species (5, 22.7%) and *Klebsiella oxytoca* (3, 13.6%). Similar studies in the West reveal a greater incidence of Coagulase-negative staphylococci and Enterococci(8).

Samples from mucocutaneous sites such as the oral cavity and perianal sites revealed a predominance of Enterobacterales, namely *Klebsiella oxytoca* (14, 35.9%), *Klebsiella pneumoniae* (7, 17.9%) and *Escherichia coli* (5, 12.8%). The proportion of MDR isolates among them was 28%. Mucositis predisposes to BSI, and similar microbiological evaluation of oral cavity samples can aid in empiric antimicrobial choice(9,10).

Among FN patients with suspected BSI, pathogens were isolated in 445 FN episodes with a 30.9% blood culture positivity rate. This is in line with earlier studies from our institute and across India(11–13).

Among the fungal isolates, Candida species were predominant (23, 95%), specifically Candida albicans (12, 52%), and were most isolated from blood (17, 73.9%) in patients with acute leukemias. Candida auris was isolated in 4 BSI episodes; however, its antimicrobial susceptibility could not be reliably assessed.

The high rates of antimicrobial resistance prevalent in India are also mirrored in our study, with the proportion of all MDR isolates being 30% and 31% MDR BSI. The alarming rates of fluoroquinolone resistance, especially Levofloxacin (47.2%), severely restrict the outpatient management of low-risk FN patients. The high rates of carbapenem resistance, especially Meropenem (42.8%), require revision of empiric antimicrobial choice to include newer Beta-Lactam – Beta-Lactamase Inhibitors (BL-BLIs). Prevalence studies from India report carbapenem resistance (CR) in over 40% of *Acinetobacter baumanii* isolates due to OXA carbapenemase. However, New Delhi Metalloproteinase (NDM) consistently remains the major carbapenemase in *Enterobacteriaceae*(14).

The emergence of MDR pathogens of concern, including CRAb, CRE, MRSA, DTR Pseudomonas, *Stenotrophomonas maltophilia* and *Candida auris*, requires meticulous infection control practices and periodic surveillance to avert outbreaks(15,16). These organisms were increasingly isolated in FN patients with acute leukemias in our study. However, owing to their prolonged hospitalization, they are overrepresented in most FN studies(17). These findings suggest the use of Cefideracol or Ceftazidime-Avibactam-Aztreonam as the empiric choice in de-escalation approaches in severely ill FN patients(18).

Our single-centre and retrospective study design limits its representativeness to Eastern India. Further research involving a multicentre cohort will provide robust aid to all antimicrobial stewardship (AMS) programs in the region and India.

Our study did not find a statistically significant association between the clinical, demographic factors or hematologic disease and risk of MDR infection. However, risk prediction scoring systems have been derived from multicentric cohorts with clinical outcomes. The risk factors associated with CR-GNB include immunocompromise, central venous catheter, chemotherapy, neutropenia, prior carbapenem exposure and colonization. (10) A similar study in India detected a CR *Enterobacteriaceae* colonization rate of 18.1% among 272 samples(19). Of the 15 faecal isolates in our study, 20% were carbapenem-resistant.

## Conclusion

Our findings exemplify the dynamic nature of pathogens in FN and the need for continuous surveillance to guide empiric antimicrobial choice. The predominance of Enterobacterales suggests a re-evaluation of antipseudomonal empiric antimicrobial therapy and the consideration of newer BL-BLIs. While the potential for failure is high in similar resource-limited settings, the rewards of efficient antimicrobial stewardship are enormous among FN patients.

## Data Availability

All data produced in the present study are available upon reasonable request to the authors

